# Body Composition, Cardiometabolic Risk Factors and Comorbidities in Psoriasis and the Effect of *HLA-C*06:02* Status: The HUNT Study, Norway

**DOI:** 10.1101/2022.10.07.22280812

**Authors:** Åshild Ø. Solvin, Vera V. Bjarkø, Laurent F. Thomas, Patricia Berrospi, Kristian Hveem, Marit Saunes, Bjørn O. Åsvold, Mari Løset

## Abstract

Psoriasis has been associated with increased adiposity measures driving systemic inflammation, which may lead to metabolic dysfunction and comorbidities. In this population-based, cross-sectional study, we used data from 56 042 individuals in the fourth wave of the Trøndelag Health Study (HUNT4), to investigate the associations between psoriasis and body composition measures assessed using bioelectrical impedance analysis, cardiometabolic risk factors, and comorbidities. Further, we investigated the associations between *HLA-C*06:02* status, a potential clinical biomarker for a distinct psoriasis endotype, and these outcomes. Psoriasis was associated with increased adiposity measures, including increased body and visceral fat, and lower levels of skeletal muscle and soft lean mass, as well as higher prevalence of cardiovascular, respiratory and endocrine disorders. *HLA-C*06:02-*positive individuals with psoriasis had lower levels of hsCRP, increased prevalence of atrial fibrillation and decreased prevalence of migraine. Our results point to altered body composition in psoriasis with increased levels of fat, and particularly metabolically active visceral fat, and provide support for a broad clinical approach to psoriatic patients in a general population.

---

Psoriasis is an immune-mediated, chronic inflammatory skin disease, affecting ∼1-2% of the global population (1). The prevalence of psoriasis differs across nations, and prevalence estimates in Norway are amongst the world’s highest, ranging from 5.8-11.4% (2, 3). Psoriasis arise from a complex interplay between genomics and environmental influences, such as infections, obesity and smoking (4, 5). Observational studies have established and suggested associations between psoriasis and a range of medical conditions including psoriatic arthritis, cardiometabolic, respiratory and endocrine disorders (6, 7). Suggested mechanisms for the associations between psoriasis and comorbidities include shared genetic risk factors, immunologic pathways, vascular endothelial dysfunction and systemic inflammation (8). Common environmental risk factors, including smoking, weight gain and physical inactivity, may also contribute (4, 5). An association between psoriasis and obesity has been established, however, most studies have utilized scales and waist measurements (9). Studies with a more detailed characterization of body composition generally have lower sample sizes (9), with the largest controlled study including 242 individuals with psoriasis. The recent awareness of increased rates of several cardiometabolic risk factors and comorbidities has led attention to closer clinical surveillance and the development of clinical guidelines (7, 10). Several observational studies are based on hospital records. Less is known about the comorbid burden of psoriasis in a general population with higher prevalence estimates of psoriasis including milder cases. In these populations, the best clinical practice is unclear.

Genome-wide association studies have identified >60 loci associated with psoriasis in populations of European descent (11). The *HLA-C*06:02* risk allele has been identified as the major genetic contributor (12). *HLA-C*06:02-*positive individuals tend to have earlier age of psoriasis onset (13) and more severe disease (12, 14). Partial or total remission of psoriasis during pregnancy has been reported in *HLA-C*06:02-*positive women (14). Further, *HLA-C*06:02* could potentially serve as a genetic biomarker of systemic treatment efficacy, as *HLA-C*06:02*-positive individuals have better response to treatment with ustekinumab and metothrexate (15) and *HLA-C*06:02*-negative individuals have a better response to adalimumab (16). A recent study found *HLA-C*06:02* status to be associated with differences in clinical phenotype and risk of comorbidities, including hypertension, psoriatic arthritis, peptic ulcer, thyroid disease and depression (17). This study also identified sex-specific effects on age of psoriasis onset and adiposity measures. *HLA-C*06:02-*positive women had earlier disease onset than *HLA-C*06:02-*positive men and *HLA-C*06:02*-negative women had higher measures of central adiposity compared to *HLA-C*06:*02-negative men (17). This makes *HLA-C*06:02* a potentially important biomarker of increased risk for more severe disease and altered comorbid burden. The study established *HLA-C*06:02-*positive individuals as a distinct disease endotype with implications for disease development, burden and treatment (17).

In this study, we aimed to assess the overall prevalence of psoriasis, and to estimate the associations of psoriasis with body composition measures assessed using bioelectrical impedance analysis, cardiometabolic risk factors, and comorbidities in a large population-based study (HUNT4) in Norway. Further we aimed to investigate the recently reported association of *HLA-C*06:02* status, anthropometric measures, comorbidities, and for the first time, investigate associations with detailed body composition measurements.

## MATERIALS AND METHODS

### Study population

The Trøndelag Health Study (HUNT) is an ongoing population-based study, which has collected data in four surveys (18, 19). We used data from HUNT4, carried out in 2017-2019 (18). All inhabitants in the Nord-Trøndelag region aged 20 years or older were invited and 56 042 (54.0%) participated. Participation included extensive questionnaires regarding lifestyle and medical history, non-fasting blood samples and clinical examinations. Participants with missing information regarding psoriasis status were excluded (n = 2669) and the population studied included 53 373 individuals. As some participants had missing information on outcomes, the number of individuals included in each analysis varies. A total of 16 434 individuals were genotyped for rs4406273, of which 931 had psoriasis. For models including genetic information, we excluded non-European and related individuals, leaving a population of 901 psoriatic individuals for analysis.

### Classification of psoriasis

Psoriasis was defined as an affirmative response to the following cluster question in HUNT4: “Have you ever had any of the following diseases?” “Psoriasis”. Participants answering no to this question were defined as individuals without psoriasis. The psoriasis question has been validated in the third wave of HUNT (HUNT3) conducted between 2006 and 2008, with a positive predictive value of 78% (2).

### Classification of comorbidities

The aforementioned cluster question also included myocardial infarction, angina, cardiac failure, atrial fibrillation, apoplexia, asthma, chronic obstructive pulmonary disorder (COPD), diabetes, hypothyroidism, hyperthyroidism, migraine, renal disease and gout. A positive response to either disease was classified as having that disease. In addition, biochemical measurements were used when available. Diabetes was defined as an affirmative answer and/or an HbA1c ≥ 48 mmol/mol. Hypothyroidism was defined as having an affirmative answer and/or a TSH >4.5 mU/L and free thyroxine <9.0 pmol/L. Hyperthyroidism was defined as an affirmative answer and/or a TSH <0.1 mU/L and free thyroxine >19 pmol/L in individuals without known hypothyroidism. Correspondingly, kidney disease was defined by an affirmative answer and/or an estimated glomerular filtration rate (eGFR) of ≤60 mL/min/1.73 m^2^.

### Clinical examination and laboratory measurements

All clinical examinations were performed by specially trained staff. Blood pressure was measured using Dinamap 845XT based on oscillometry. The average value of the second and third measurement was used. Height and weight were measured with the participants wearing light clothes and no shoes. Body mass index (BMI) was calculated by dividing weight in kilograms by height in meters squared (kg/m^2^). Body composition of all participants was assessed using bioelectrical impedance analysis, on an *InBody 770-*instrument (Cerritos, CA, USA). Body composition parameters reported in kilograms (total body fat, skeletal muscle mass, soft lean mass and fat free mass) were reported as a percentage of total body weight. Analyses of non-fasting blood samples were carried out using Architect cSystems ci8200 (Abbott Diagnostics, Longford Ireland).

### Genotyping

Genotyping was performed using a custom Illumina HumanCoreExome array (UM HUNT Biobank v.2.0) on 18 722 individuals (20). Ancestry was predicted using principal components of ancestry projected onto the 1000 Genomes Project. Ancestry principal components were generated, and eigen-values were estimated (Supplementary figs. 1 and 2). The single nucleotide polymorphism (SNP) rs4406273 was used as a proxy-SNP for *HLA-C*06:02* status, which has shown excellent performance (Matthews correlation coefficient 0.965-1.000) (21). Individuals with psoriasis were classified as either *HLA-C*06:02-*positive (one or two copies of the rs4406273-A allele) or *HLA-C*06:02-*negative (no copies).

### Statistical analyses

All descriptive variables (except sex) are reported standardized for age applying Stata’s analytic weights option, where observations are weighted inversely proportional to the variance of the observation. Means and 95% confidence intervals (95% CI) were reported for continuous variables and counts and percentages were reported for categorical variables. A generalized linear model was applied to estimate prevalence ratios (PR) for the different outcomes in the psoriatic and non-psoriatic groups. In cases where the model did not converge, we calculated differences in proportion based on predicted mean probabilities from a logistic regression model using Stata’s adjrr command. To adjust for potential confounding in the models, covariates were chosen based on *a priori* knowledge on disease mechanisms. Two models were constructed; one adjusting for age, sex and BMI and another, fully adjusted model, also adjusting for smoking status, education and alcohol consumption. Subsequently, a sub-analysis was performed among psoriatic individuals, using *HLA-C*06:02* status as the exposure and self-reported disease as outcome. The model was adjusted for potential mediators and confounders; age, sex, BMI, smoking status, disease duration and cryptic population structure by four principal components. Precision was assessed by 95% CI. All statistical analyses were conducted using Stata MP v.17.0 (StataCorp, College Station, TX, USA).

### Ethics

The study was approved by the Regional Committee for Medical and Health Research Ethics in Central Norway (REK Reference number: 27420), and the Norwegian Data Protection Authority. All HUNT participants gave written informed consent.

## RESULTS

### Characteristics of the study population

Demographic details of the 53 373 participants are provided in Table I. A total of 3 535 individuals (6.6%) reported psoriasis. Individuals with psoriasis were older, more frequently smokers, and more often *HLA-C*06:02*-positive than individuals without psoriasis (Table I). The frequency of *HLA-C*06:02-*positivity was 26.6% among individuals with psoriasis and 11.7% among individuals without psoriasis.

**Table I.**
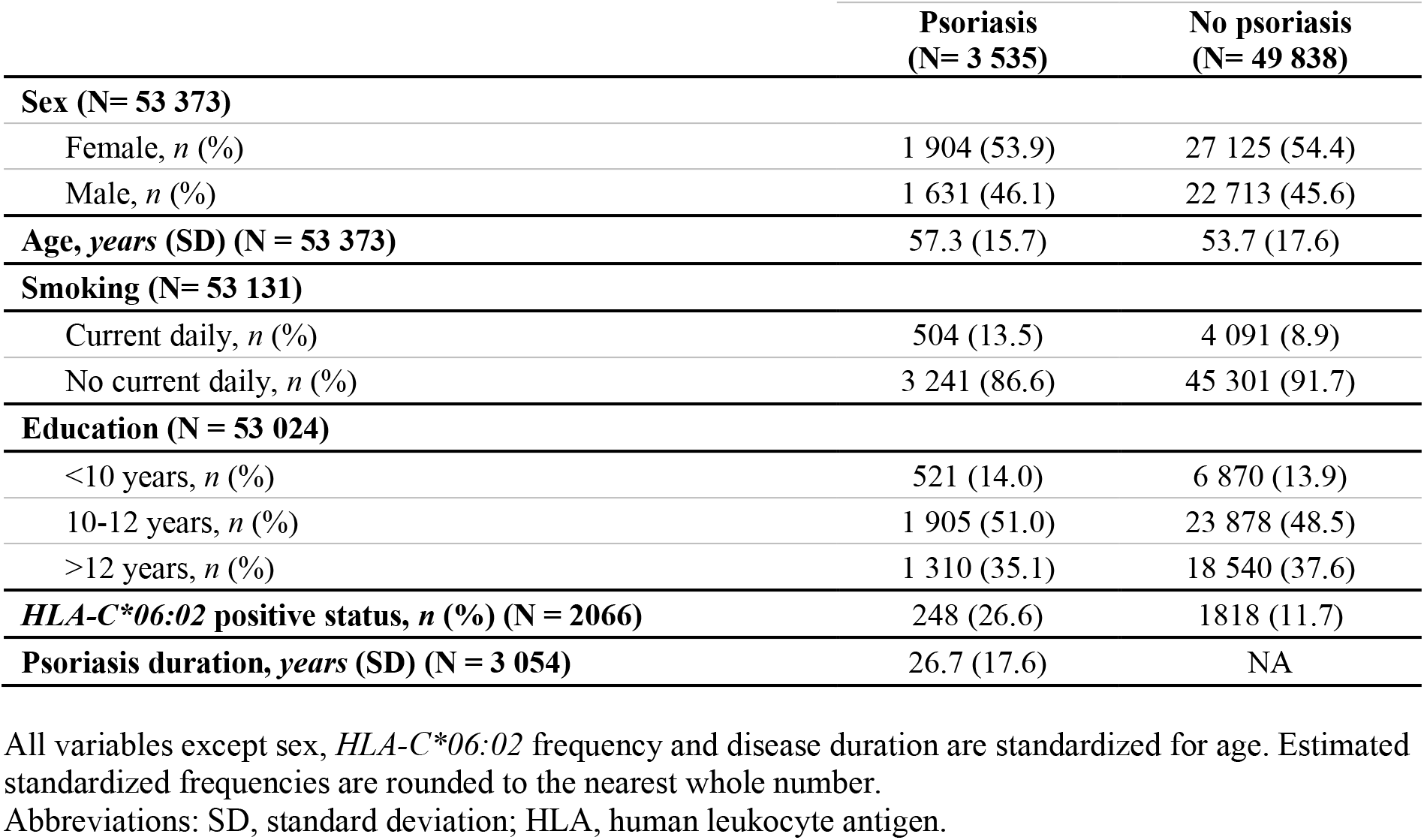
Demographic characteristics of the study population as stratified by psoriasis.

|  | **Psoriasis**<br>**(N= 3 535)** | **No psoriasis**<br>**(N= 49 838)** |
| --- | --- | --- |
| **Sex (N= 53 373)** |  |  |
| Female, *n* (%) | 1 904 (53.9) | 27 125 (54.4) |
| Male, *n* (%) | 1 631 (46.1) | 22 713 (45.6) |
| **Age, years (SD) (N = 53 373)** | 57.3 (15.7) | 53.7 (17.6) |
| **Smoking (N= 53 131)** |  |  |
| Current daily, *n* (%) | 504 (13.5) | 4 091 (8.9) |
| No current daily, *n* (%) | 3 241 (86.6) | 45 301 (91.7) |
| **Education (N = 53 024)** |  |  |
| <10 years, *n* (%) | 521 (14.0) | 6 870 (13.9) |
| 10-12 years, *n* (%) | 1 905 (51.0) | 23 878 (48.5) |
| >12 years, *n* (%) | 1 310 (35.1) | 18 540 (37.6) |
| **HLA-C\*06:02 positive status, *n* (%) (N = 2066)** | 248 (26.6) | 1818 (11.7) |
| **Psoriasis duration, years (SD) (N = 3 054)** | 26.7 (17.6) | NA |

All variables except sex, *HLA-C\*06:02* frequency and disease duration are standardized for age. Estimated standardized frequencies are rounded to the nearest whole number.
Abbreviations: SD, standard deviation; HLA, human leukocyte antigen.

### Body composition parameters and cardiometabolic risk factors

Individuals with psoriasis had higher BMI and waist circumference compared to individuals without psoriasis (Table II). They also had higher total body fat (33.3% vs. 31.5%) and visceral fat (138.6 vs. 125.5 cm^2^), as well as lower levels of skeletal muscle mass (36.8% vs. 37.8%), soft lean mass (62.9% vs. 64.5%) and fat free mass (66.7% vs. 68.5%) (Table II). Females and males contributed equally to these differences (Supplementary table SII). Individuals with psoriasis had higher levels of systolic blood pressure (132.0 vs. 131.1 mmHg), triglycerides (1.8 vs. 1.7 mmol/L), HbA1c (36.4 vs. 35.4 mmol/mol) and high sensitivity CRP (hsCRP) (3.1 vs. 2.7 mg/L) compared to individuals without psoriasis. For systolic blood pressure, this difference was greater in females than in males. *HLA-C*06:02-* negative psoriatic individuals had a tendency towards higher levels of percentage body fat and visceral fat compared to *HLA-C*06:02-*positive, with no apparent difference between the sexes (Supplementary table SII). *HLA-C*06:02-*positive psoriatic individuals had lower levels of hsCRP compared to *HLA-C*06:02-*negative psoriatic individuals (2.6 vs. 3.3 mg/L) and this difference appeared to sex specific, with the female contribution dominating (Supplementary table SII).

**Table II.**
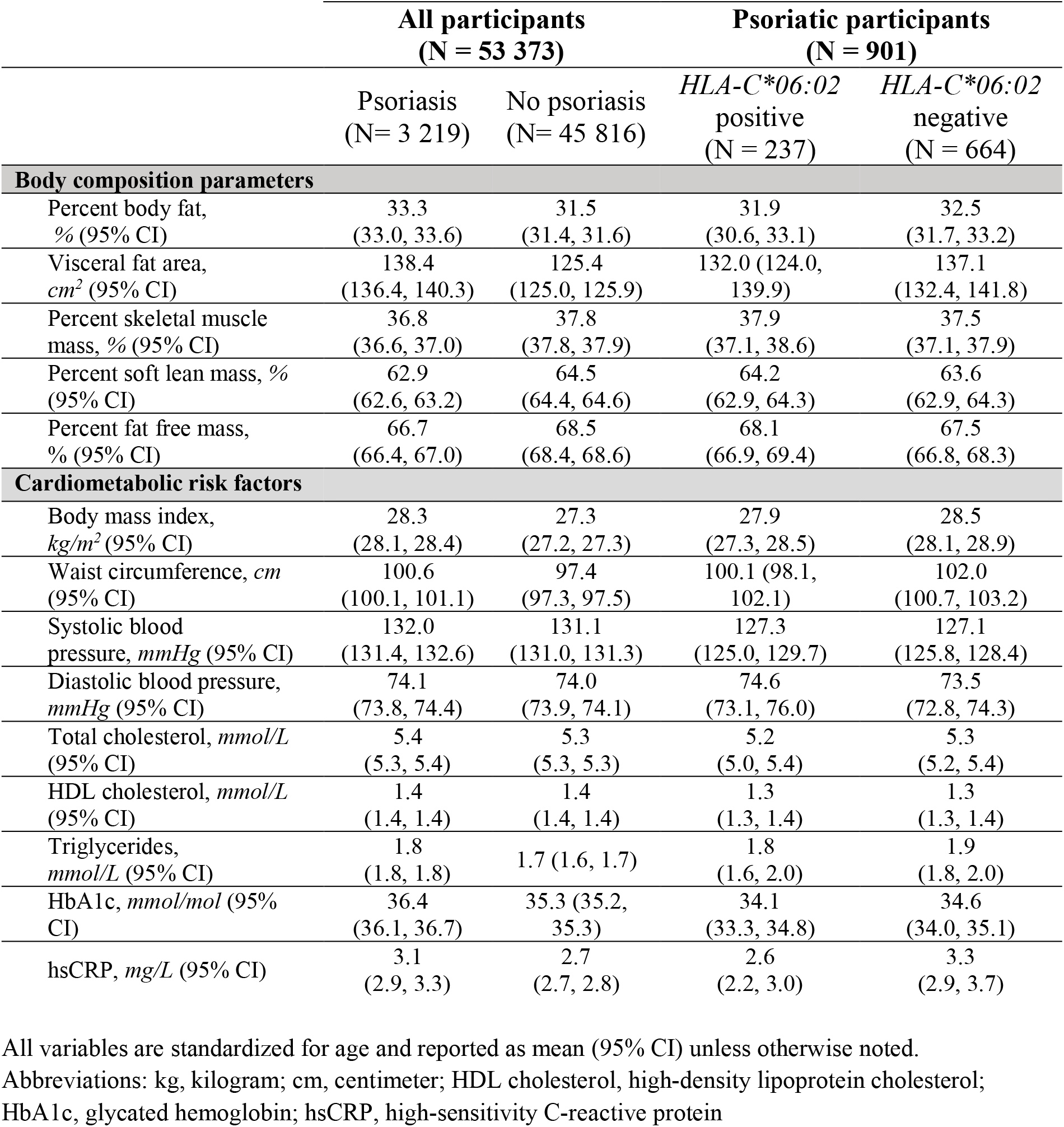
Body composition measures and cardiometabolic risk factors in individuals with and without psoriasis.

|  | **All participants**<br>**(N = 53 373)** |  | **Psoriatic participants**<br>**(N = 901)** |  |
| --- | --- | --- | --- | --- |
|  | Psoriasis<br>(N= 3 219) | No psoriasis<br>(N= 45 816) | *HLA-C\*06:02*<br>positive<br>(N = 237) | *HLA-C\*06:02*<br>negative<br>(N = 664) |
| **Body composition parameters** |  |  |  |  |
| Percent body fat,<br>% (95% CI) | 33.3<br>(33.0, 33.6) | 31.5<br>(31.4, 31.6) | 31.9<br>(30.6, 33.1) | 32.5<br>(31.7, 33.2) |
| Visceral fat area,<br>*cm*<sup>2</sup> (95% CI) | 138.4<br>(136.4, 140.3) | 125.4<br>(125.0, 125.9) | 132.0 (124.0,<br>139.9) | 137.1<br>(132.4, 141.8) |
| Percent skeletal muscle<br>mass, % (95% CI) | 36.8<br>(36.6, 37.0) | 37.8<br>(37.8, 37.9) | 37.9<br>(37.1, 38.6) | 37.5<br>(37.1, 37.9) |
| Percent soft lean mass, %<br>(95% CI) | 62.9<br>(62.6, 63.2) | 64.5<br>(64.4, 64.6) | 64.2<br>(62.9, 64.3) | 63.6<br>(62.9, 64.3) |
| Percent fat free mass,<br>% (95% CI) | 66.7<br>(66.4, 67.0) | 68.5<br>(68.4, 68.6) | 68.1<br>(66.9, 69.4) | 67.5<br>(66.8, 68.3) |
| **Cardiometabolic risk factors** |  |  |  |  |
| Body mass index,<br>*kg/m*<sup>2</sup> (95% CI) | 28.3<br>(28.1, 28.4) | 27.3<br>(27.2, 27.3) | 27.9<br>(27.3, 28.5) | 28.5<br>(28.1, 28.9) |
| Waist circumference, *cm*<br>(95% CI) | 100.6<br>(100.1, 101.1) | 97.4<br>(97.3, 97.5) | 100.1 (98.1,<br>102.1) | 102.0<br>(100.7, 103.2) |
| Systolic blood<br>pressure, *mmHg* (95% CI) | 132.0<br>(131.4, 132.6) | 131.1<br>(131.0, 131.3) | 127.3<br>(125.0, 129.7) | 127.1<br>(125.8, 128.4) |
| Diastolic blood pressure,<br>*mmHg* (95% CI) | 74.1<br>(73.8, 74.4) | 74.0<br>(73.9, 74.1) | 74.6<br>(73.1, 76.0) | 73.5<br>(72.8, 74.3) |
| Total cholesterol, *mmol/L*<br>(95% CI) | 5.4<br>(5.3, 5.4) | 5.3<br>(5.3, 5.3) | 5.2<br>(5.0, 5.4) | 5.3<br>(5.2, 5.4) |
| HDL cholesterol, *mmol/L*<br>(95% CI) | 1.4<br>(1.4, 1.4) | 1.4<br>(1.4, 1.4) | 1.3<br>(1.3, 1.4) | 1.3<br>(1.3, 1.4) |
| Triglycerides,<br>*mmol/L* (95% CI) | 1.8<br>(1.8, 1.8) | 1.7 (1.6, 1.7) | 1.8<br>(1.6, 2.0) | 1.9<br>(1.8, 2.0) |
| HbA1c, *mmol/mol* (95%<br>CI) | 36.4<br>(36.1, 36.7) | 35.3 (35.2,<br>35.3) | 34.1<br>(33.3, 34.8) | 34.6<br>(34.0, 35.1) |
| hsCRP, *mg/L* (95% CI) | 3.1<br>(2.9, 3.3) | 2.7<br>(2.7, 2.8) | 2.6<br>(2.2, 3.0) | 3.3<br>(2.9, 3.7) |

All variables are standardized for age and reported as mean (95% CI) unless otherwise noted.
Abbreviations: kg, kilogram; cm, centimeter; HDL cholesterol, high-density lipoprotein cholesterol;
HbA1c, glycated hemoglobin; hsCRP, high-sensitivity C-reactive protein

### Comorbidities

The prevalence ratios for all comorbidities were adjusted for age and gender, and in the fully adjusted model for age, gender, BMI, smoking and education (Table III). Individuals with psoriasis had higher prevalence of cardiovascular disease, including myocardial infarction (adjusted prevalence ratio (aPR) 1.54) angina pectoris (aPR 1.55), heart failure (aPR 1.56), atrial fibrillation (aPR 1.41) and apoplexia (aPR 1.63) (Table III). The prevalence of asthma (aPR 1.51), COPD (aPR 1.89) diabetes (aPR 1.61), hypothyroidism (aPR 1.32) and hyperthyroidism (aPR 1.58) were higher among individuals with psoriasis. Moreover, the prevalence of migraine (aPR 1.41), renal disease (aPR 1.63) and gout (aPR 1.83) were higher in individuals with psoriasis (Table III). Additional adjustment for BMI, education and smoking reduced the strength of the associations, but they all remained significant.

**Table III.** Prevalence ratio (PR) for comorbidities in individuals with psoriasis compared to those without psoriasis.

|  | **Psoriasis**<br>**(N= 3535)** | **No psoriasis**<br>**(N= 49 838)** | **Adjusted PR**<br>**(95% CI)<sup>a</sup>** | **Fully adjusted PR**<br>**(95% CI)<sup>b</sup>** |
| --- | --- | --- | --- | --- |
| **Cardiovascular disease** |  |  |  |  |
| Myocardial infarction, *n* (%)<br>(N = 52 995) | 202 (6.0) | 1664 (3.3) | 1.54<br>(1.35, 1.76) | 1.44<br>(1.26, 1.66) |
| Angina pectoris, *n* (%)<br>(N = 52 811) | 155 (4.6) | 1308 (2.7) | 1.55<br>(1.33, 1.82) | 1.53<br>(1.30, 1.80) |
| Heart failure, *n* (%)<br>(N = 52 779) | 90 (2.7) | 762 (1.5) | 1.56<br>(1.27, 1.93) | 1.54<br>(1.24, 1.92) |
| Atrial fibrillation, *n* (%)<br>(N = 52 486) | 259 (7.8) | 2430 (4.9) | 1.41<br>(1.25, 1.59) | 1.35<br>(1.19, 1.52) |
| Apoplexia, *n* (%)<br>(N = 52 780) | 185 (5.5) | 1506 (3.1) | 1.63<br>(1.41, 1.88) | 1.56<br>(1.34, 1.81) |
| **Respiratory disease** |  |  |  |  |
| Asthma, *n* (%)<br>(N = 52 915) | 552 (16.4) | 5612 (11.3) | 1.51<br>(1.40, 1.64) | 1.45<br>(1.34, 1.58) |
| Chronic obstructive<br>pulmonary disease, *n* (%)<br>(N = 52 926) | 175 (5.2) | 1227 (2.5) | 1.89<br>(1.63, 2.20) | 1.72<br>(1.47, 2.01) |
| **Endocrine disease** |  |  |  |  |
| Diabetes, *n* (%) (N = 53 243) | 336 (9.6) | 2699 (5.4) | 1.61<br>(1.45, 1.79) | 1.49<br>(1.31, 1.69) |
| Hypothyroidism, *n* (%)<br>(N = 52 499) | 293 (8.8) | 3072 (6.3) | 1.32<br>(1.18, 1.47) | 1.27<br>(1.14, 1.43) |
| Hyperthyroidism, *n* (%)<br>(N = 52 275) | 98 (3.0) | 887 (1.8) | 1.58<br>(1.28, 1.93) | 1.54<br>(1.25, 1.89) |
| **Others** |  |  |  |  |
| Migraine, *n* (%) (N = 52 930) | 707 (21.0) | 7622 (15.4) | 1.41<br>(1.31, 1.50) | 1.39<br>(1.30, 1.49) |
| Renal disease (Non-UTI), *n*<br>(%) (N = 52 909) | 188 (5.6) | 1584 (3.2) | 1.63<br>(1.41, 1.89) | 1.53<br>(1.31, 1.78) |
| Gout, *n* (%) (N = 52 791) | 246 (7.3) | 1778 (3.6) | 1.83<br>(1.61, 2.07) | 1.63<br>(1.44, 1.85) |

<sup>a</sup> Adjusted for age and sex
<sup>b</sup> Adjusted for age, sex, BMI, education and smoking
Abbreviations: CI, confidence interval; UTI, urinary tract infection; BMI, body mass index

*HLA-C*06:02*-positive individuals had higher prevalence of atrial fibrillation (aPR 3.96,) and slightly lower prevalence of migraine (aPR 0.73) compared to *HLA-C*06:02*-negative individuals (Table IV).

**TTable IV.**
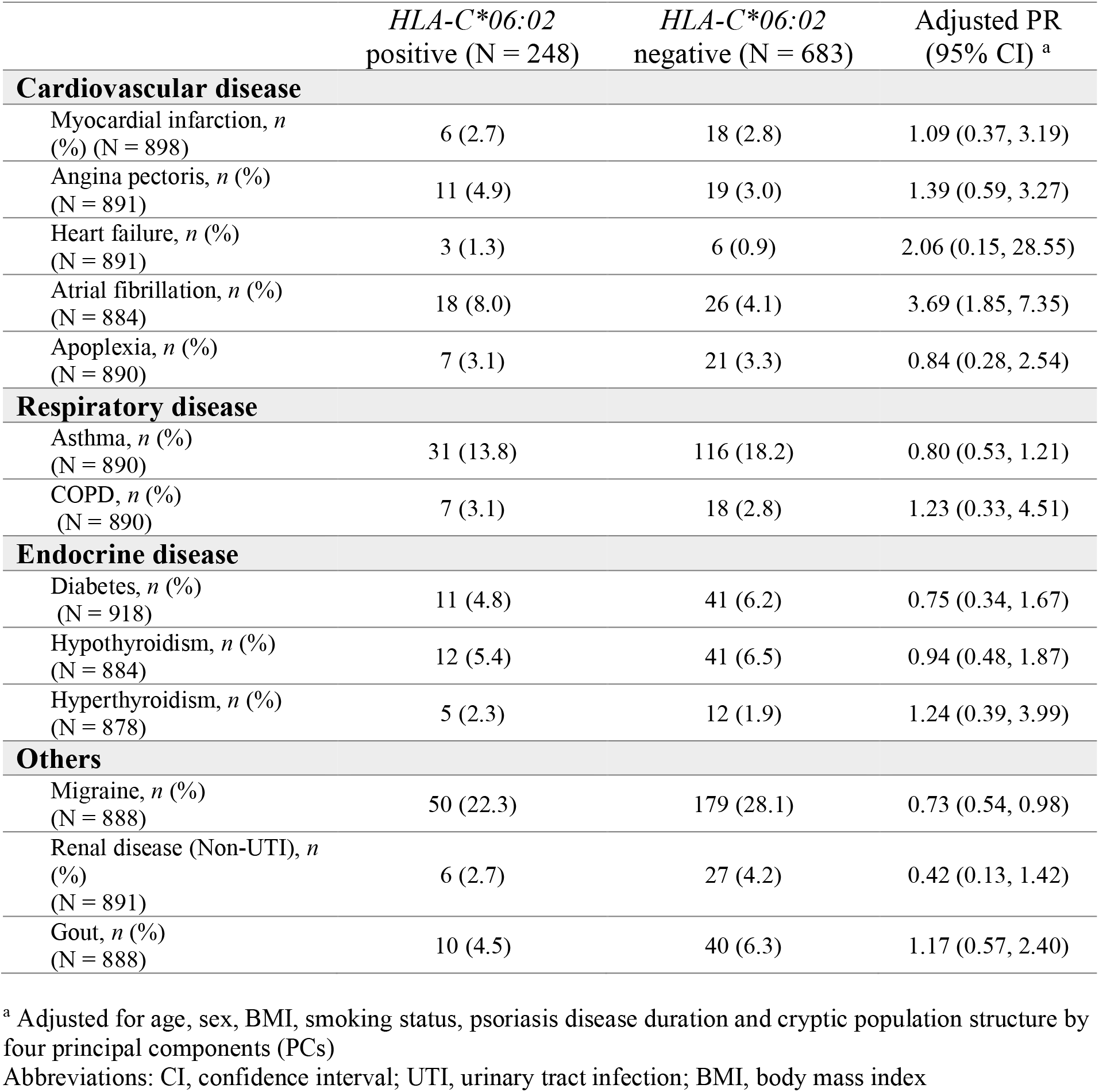
Prevalence ratio (PR) for comorbidities in *HLA-C*06:02*-positive compared with *HLA-C*06:02*-negative individuals with psoriasis.

## DISCUSSION

In this population-based cross-sectional study, we found a cumulative prevalence for psoriasis of 6.6%. To the best of our knowledge, we have performed the largest study to date evaluating detailed body composition measurements in psoriasis, and confirm previous associations with increased adiposity measures, in particular, metabolically active visceral fat. Our results support previously described associations between psoriasis and increased levels of cardiometabolic risk factors and higher prevalence of a wide range of other medical conditions in individuals with psoriasis. We extend previous studies by investigating associations between detailed body composition measurements, and *HLA-C*06:02* status among individuals with psoriasis. *HLA-C*06:02*-positive individuals had lower percentages of total body fat and visceral fat, lower levels of hsCRP, increased prevalence of atrial fibrillation, and decreased prevalence of migraine.

We found a self-reported prevalence of psoriasis in HUNT4 of 6.6%, which is higher than what has been reported for western Europe (1.9%) (1), but lower than the 11.4% reported in another Norwegian population further north (3). Our results support previous findings of higher rates of psoriasis at higher latitudes (4). Further, the high prevalence in this general population may partly be explained by the identification of milder forms of psoriasis not typically included in hospital-based studies (2).

Adipose tissue can be viewed as an endocrine organ with the capability to induce an inflammatory state through secretion of proinflammatory cytokines including T helper 17 cytokines (9). Inflammatory mediators are suggested to take part in driving the development and worsening of psoriasis and comorbidities (8, 9). Genetically influenced higher BMI has been shown to causally increase the risk of developing psoriasis (5). As adiposity is a potential modifiable risk factor for both psoriasis and several comorbidities, there are important implications of increasing our knowledge of the underlying mechanisms and to evaluate the different types of adiposity independently. Body composition evaluation is superior to the most used metric to classify obesity, namely BMI, as it provides information on lean mass and fat mass distribution, valuable gauges of metabolic health. In addition, body composition assessment using bioelectrical impedance is more sensitive at identifying obesity than BMI (22). Studies have examined the associations between psoriasis and a detailed body compositions, but the results are unclear, potentially due to sample sizes (≤252 individuals with psoriasis) and the use of different assessment modalities across studies (9). Our body composition analyses included 3219 individuals with psoriasis and 45 816 individuals without psoriasis, and we observed that individuals with psoriasis tend to have more body fat, particularly visceral fat, than those without psoriasis. This is corroborated by findings by positron-emission tomography (PET)/computed tomography (CT) in 77 individuals with psoriasis (23) and in a recent systematic review (9).

Visceral adiposity and associated low-grade inflammation have been implicated in the development of a wide range of diseases, including cardiovascular, autoinflammatory, kidney and liver disease (8). *HLA-C*06:02*-positive psoriatic individuals had lower levels of total body fat and visceral fat than *HLA-C*06:02-*negative psoriatic individuals. This is in line with previous studies investigating the association between *HLA-C*06:02* status and obesity measured by BMI and waist circumference (17). Notably, we also found that *HLA-C*06:02-* negative women in particular have increased levels of hsCRP, a well-known biomarker of cardiovascular risk. This is in line with previous studies which have found higher prevalence of cardiometabolic risk in women (24), and in particular, *HLA-C*06:02-*negative women (17). We found that individuals with psoriasis had lower levels of skeletal muscle and soft lean mass by total body weight compared to individuals without psoriasis. Still, despite individuals with psoriasis being on average 2.74 kg heavier than individuals without psoriasis, only 9 g (0.33%) of this difference was made up of muscle mass. Sustained inflammation is shown to give loss of muscle mass and strength (9), and may partly explain the observed low increase in muscle mass relative to increase in body weight.

In recent years, increasing attention has been given towards the association between psoriasis and a wide range of comorbidities. This has proven important to improve quality of life and to reduce the risk of overall mortality among psoriasis patients (25). Aggressive psoriasis treatments may modify cardiovascular mortality rates (26). Further, comorbidities affect the likelihood of achieving systemic treatment response (27). Obesity is shown to give poorer outcomes in biologic treatment efficacy, and a recent US-based study expanded this knowledge to include also other metabolic comorbid conditions at six months (28). To provide specific clinical guidelines for appropriate follow-up, the American Academy of Dermatology (AAD) and the National Psoriasis Foundation (NPF) published Joint Guidelines with recommendations for the management and treatment of comorbidities associated with psoriasis (7). In 2021, the European Academy of Dermatology followed suit (10). However, it has been unclear if knowledge acquired in a hospital-based setting can be applied to a general population with higher prevalence estimates of psoriasis. In the present study, we have provided evidence and replicated associations between psoriasis and a range of cardiovascular diseases (myocardial infarction, angina, heart failure, atrial fibrillation, and apoplexia) (29–31), respiratory diseases (asthma and COPD) (32, 33) and endocrine diseases (diabetes, hypothyroidism, and hyperthyroidism) (34, 35), as well as kidney disease (36), migraine (37) and gout (38). Direct comparison of effect sizes between different studies is difficult due to differences in methodology. However, many studies are based on data on hospitalized patients and may not accurately reflect the burden of disease in a general population. In the present study, associations remain significant after adjusting for age, sex, BMI, education and smoking, however, residual confounding may still occur.

Personalized approaches to treatment and management of psoriasis require the identification of robust biomarkers. Genetic markers may predict certain comorbidities and the effectiveness of treatment, improving patient outcomes. A previous study among psoriasis patients demonstrated that 200 genetic markers were predictive for development of psoriatic arthritis development (area under the receiver operator curve, AUROC = 0.82) (39) and genetic biomarkers can be used to predict treatment efficacy and safety (40). When investigating potential associations between *HLA-C*06:02* status and comorbidities in the psoriasis population, we found increased prevalence of atrial fibrillation and decreased prevalence of migraine in *HLA-C*06:02*-positive individuals. It should be noted that the confidence intervals are wide, and results should therefore be interpreted with caution. In a study of individuals with psoriasis by Douroudis et al., *HLA-C*06:02-*negativity was associated with higher prevalence of cardiovascular disease and thyroid disease (17), findings which were not replicated in the present study. In the aforementioned study, no significant association between *HLA-C*06:02* status and type 2 diabetes or asthma was found (17). In the present study, the samples sizes were too small to meaningfully discuss sex-specific effects of *HLA-C*06:02* status on prevalence of comorbidities, as these analyses yielded very wide confidence intervals (Supplementary table IV).

A major strength of this study is the population-based design with a high participation rate of 54.0%. As Norway has one of the world’s highest reported prevalences of psoriasis, studies on Norwegian populations may be of particular interest. To the best of our knowledge, the study is also by far the largest study to date reporting detailed body composition measures in individuals with psoriasis. The use of genetic data in relation to comorbidities contributes to the novelty of this paper as this aspect has sparsely been studied previously. This study did have several limitations. Firstly, we did not investigate subgroups of psoriasis regarding clinical subtype nor severity. In addition, most disease outcomes were self-reported, introducing the possibility of misclassification. However, the psoriasis question in HUNT has previously been validated by experienced dermatologists, with an overall positive predictive value of 78% (2), indicating that self-reported psoriasis in HUNT is a reliable source of data for research purposes. In addition, several of the self-reported variables, including migraine, diabetes, and atrial fibrillation has been validated (18). The present study is at risk of ascertainment bias, wherein individuals with psoriasis are screened for other diseases more frequently than controls due to more frequent contacts with health care services. This may introduce false positive associations between psoriasis and comorbidities.

In conclusion, this large population-based cross-sectional study shows persistently high prevalence of psoriasis in Norway. We have identified increased levels of total body fat and visceral fat, as well as lower levels of skeletal muscle, and soft lean mass in individuals with psoriasis. In addition, we replicated previous findings indicating that *HLA-C*06:02-*negative individuals, and women in particular, have an increased cardiometabolic risk burden. We also replicated several associations between psoriasis, cardiometabolic risk factors and comorbidities, and identified an increased prevalence of atrial fibrillation and decreased prevalence of migraine in *HLA-C*06:02-*positive individuals. Our results support the importance of addressing psoriasis as a systemic disease that requires holistic and multidisciplinary care. Special attention should be made towards individuals with psoriasis and obesity and our results highlights the importance of identification of cardiometabolic risk factors and prevention of comorbidities both for the hospital-based dermatologist, as well as for the general practitioner treating these patients in a general population.

## Data Availability

Researchers associated with Norwegian research institutes can apply for the use of HUNT data and samples with approval by the Regional Committee for Medical and Health Research Ethics. Researchers from other countries may apply if collaborating with a Norwegian Principal Investigator. Information for data access can be found at https://www.ntnu.edu/hunt/data. The HUNT variables are available for browsing on the HUNT databank at https://hunt-db.medisin.ntnu.no/hunt-db/. Data linkages between HUNT and health registries require that the principal investigator has obtained project-specific approval for such linkage from the Regional Committee for Medical and Health Research Ethics, Norway and each registry owner.

## Acknowledgements

We are very grateful to all HUNT participants for donating their time, samples and information to help others and we thank the personnel at the screening surveys. The Trøndelag Health Study (The HUNT Study) is a collaboration between HUNT Research Center (Faculty of Medicine and Health Sciences, NTNU - Norwegian University of Science and Technology), Trøndelag County Council, Central Norway Regional Health Authority, and the Norwegian Institute of Public Health. The genetic investigations of the HUNT Study is a collaboration between researchers from the K.G. Jebsen Center for Genetic Epidemiology and University of Michigan Medical School, and the University of Michigan School of Public Health. The K.G. Jebsen Center for Genetic Epidemiology is financed by Stiftelsen Kristian Gerhard Jebsen; Faculty of Medicine and Health Sciences, NTNU, Norway. ML is supported by grants from the Liaison Committee for education, research and innovation in Central Norway. We thank Marita Jenssen for her suggestions on this manuscript.

## Notes

### Competing Interest Statement

The authors have declared no competing interest.

### Author Declarations

The Regional Committee for Medical and Health Research Ethics of Central Norway gave ethical approval for this work (REK Reference number: 27420). All participants signed informed consent.

### Summary of Updates

Methodology updated to provide accurate company names of instruments used in the study and correct a possible error in the description of the measurement of waist circumference. Results section on comorbidities updated to correct a misinterpretation of data. Updated titles of tables and supplementary tables.

## REFERENCES

1. Parisi R, Iskandar IYK, Kontopantelis E, Augustin M, Griffiths CEM, Ashcroft DM. National, regional, and worldwide epidemiology of psoriasis: systematic analysis and modelling study. BMJ. 2020;369:m1590.

2. Modalsli EH, Snekvik I, Asvold BO, Romundstad PR, Naldi L, Saunes M. Validity of Self-Reported Psoriasis in a General Population: The HUNT Study, Norway. The Journal of investigative dermatology. 2016;136(1):323–5.

3. Danielsen K, Olsen AO, Wilsgaard T, Furberg AS. Is the prevalence of psoriasis increasing? A 30-year follow-up of a population-based cohort. The British journal of dermatology. 2013;168(6):1303–10.

4. Griffiths CEM, Armstrong AW, Gudjonsson JE, Barker J. Psoriasis. Lancet (London, England). 2021;397(10281):1301–15.

5. Budu-Aggrey A, Brumpton B, Tyrrell J, Watkins S, Modalsli EH, Celis-Morales C, et al. Evidence of a causal relationship between body mass index and psoriasis: A mendelian randomization study. PLoS Med. 2019;16(1):e1002739.

6. Takeshita J, Grewal S, Langan SM, Mehta NN, Ogdie A, Van Voorhees AS, et al. Psoriasis and comorbid diseases: Epidemiology. J Am Acad Dermatol. 2017;76(3):377–90.

7. Elmets CA, Leonardi CL, Davis DMR, Gelfand JM, Lichten J, Mehta NN, et al. Joint AAD-NPF guidelines of care for the management and treatment of psoriasis with awareness and attention to comorbidities. J Am Acad Dermatol. 2019;80(4):1073–113.

8. Hao Y, Zhu Y-j, Zou S, Zhou P, Hu Y-w, Zhao Q-x, et al. Metabolic Syndrome and Psoriasis: Mechanisms and Future Directions. Front Immunol. 2021;12.

9. Blake T, Gullick NJ, Hutchinson CE, Barber TM. Psoriatic disease and body composition: A systematic review and narrative synthesis. PloS one. 2020;15(8):e0237598–e.

10. Nast A, Smith C, Spuls PI, Avila Valle G, Bata-Csörgö Z, Boonen H, et al. EuroGuiDerm Guideline on the systemic treatment of Psoriasis vulgaris – Part 2: specific clinical and comorbid situations. Journal of the European Academy of Dermatology and Venereology. 2021;35(2):281–317.

11. Tsoi LC, Stuart PE, Tian C, Gudjonsson JE, Das S, Zawistowski M, et al. Large scale meta-analysis characterizes genetic architecture for common psoriasis associated variants. Nature communications. 2017;8(1):1–8.

12. Fan X, Yang S, Sun LD, Liang YH, Gao M, Zhang KY, et al. Comparison of clinical features of HLA-Cw*0602-positive and -negative psoriasis patients in a Han Chinese population. Acta Derm Venereol. 2007;87(4):335–40.

13. Henseler T, Christophers E. Psoriasis of early and late onset: characterization of two types of psoriasis vulgaris. J Am Acad Dermatol. 1985;13(3):450–6.

14. Gudjonsson JE, Karason A, Hjaltey Runarsdottir E, Antonsdottir AA, Hauksson VB, Jónsson HH, et al. Distinct Clinical Differences Between HLA-Cw*0602 Positive and Negative Psoriasis Patients – An Analysis of 1019 HLA-C- and HLA-B-Typed Patients. Journal of Investigative Dermatology. 2006;126(4):740–5.

15. Chen L, Tsai TF. HLA-Cw6 and psoriasis. The British journal of dermatology. 2018;178(4):854–62.

16. Dand N, Duckworth M, Baudry D, Russell A, Curtis CJ, Lee SH, et al. HLA-C*06:02 genotype is a predictive biomarker of biologic treatment response in psoriasis. The Journal of allergy and clinical immunology. 2019;143(6):2120–30.

17. Douroudis K, Ramessur R, Barbosa IA, Baudry D, Duckworth M, Angit C, et al. Differences in Clinical Features and Comorbid Burden between HLA-C*06:02 Carrier Groups in >9,000 People with Psoriasis. The Journal of investigative dermatology. 2022;142(6):1617–28.e10.

18. Åsvold BO, Langhammer A, Rehn TA, Kjelvik G, Grøntvedt TV, Sørgjerd EP, et al. Cohort Profile Update: The HUNT Study, Norway. Int J Epidemiol. 2022.

19. Krokstad S, Langhammer A, Hveem K, Holmen TL, Midthjell K, Stene TR, et al. Cohort Profile: the HUNT Study, Norway. Int J Epidemiol. 2013;42(4):968–77.

20. Brumpton BM, Graham S, Surakka I, Skogholt AH, Løset M, Fritsche LG, et al. The HUNT Study: a population-based cohort for genetic research. medRxiv. 2021:2021.12.23.21268305.

21. Stuart PE, Tejasvi T, Shaiq PA, Kullavanijaya P, Qamar R, Raja GK, et al. A Single SNP Surrogate for Genotyping HLA-C*06:02 in Diverse Populations. Journal of Investigative Dermatology. 2015;135(4):1177–80.

22. Galluzzo M, Talamonti M, Perino F, Servoli S, Giordano D, Chimenti S, et al. Bioelectrical impedance analysis to define an excess of body fat: evaluation in patients with psoriasis. J Dermatolog Treat. 2017;28(4):299–303.

23. Rivers JP, Powell-Wiley TM, Dey AK, Rodante JA, Chung JH, Joshi AA, et al. Visceral Adiposity in Psoriasis is Associated With Vascular Inflammation by (18)F-Fluorodeoxyglucose Positron-Emission Tomography/Computed Tomography Beyond Cardiometabolic Disease Risk Factors in an Observational Cohort Study. JACC Cardiovasc Imaging. 2018;11(2 Pt 2):349–57.

24. Guillet C, Seeli C, Nina M, Maul LV, Maul J-T. The impact of gender and sex in psoriasis: What to be aware of when treating women with psoriasis. International Journal of Women’s Dermatology. 2022;8(2).

25. Dhana A, Yen H, Yen H, Cho E. All-cause and cause-specific mortality in psoriasis: A systematic review and meta-analysis. J Am Acad Dermatol. 2019;80(5):1332–43.

26. Yang ZS, Lin NN, Li L, Li Y. The Effect of TNF Inhibitors on Cardiovascular Events in Psoriasis and Psoriatic Arthritis: an Updated Meta-Analysis. Clin Rev Allergy Immunol. 2016;51(2):240–7.

27. Enos CW, Ramos VL, McLean RR, Lin TC, Foster N, Dube B, et al. Cardiometabolic multimorbidity is common among patients with psoriasis and is associated with poorer outcomes compared to those without comorbidity. J Dermatolog Treat. 2022:1–8.

28. Enos CW, Ramos VL, McLean RR, Lin T-C, Foster N, Dube B, et al. Comorbid obesity and history of diabetes are independently associated with poorer treatment response to biologics at 6 months: A prospective analysis in Corrona Psoriasis Registry. Journal of the American Academy of Dermatology. 2022;86(1):68–76.

29. Armstrong EJ, Harskamp CT, Armstrong AW. Psoriasis and major adverse cardiovascular events: a systematic review and meta-analysis of observational studies. Journal of the American Heart Association. 2013;2(2):e000062–e.

30. Khalid U, Ahlehoff O, Gislason GH, Kristensen SL, Skov L, Torp-Pedersen C, et al. Psoriasis and risk of heart failure: a nationwide cohort study. Eur J Heart Fail. 2014;16(7):743–8.

31. Ungprasert P, Srivali N, Kittanamongkolchai W. Psoriasis and risk of incident atrial fibrillation: A systematic review and meta-analysis. Indian J Dermatol Venereol Leprol. 2016;82(5):489–97.

32. Ungprasert P, Srivali N, Thongprayoon C. Association between psoriasis and chronic obstructive pulmonary disease: A systematic review and meta-analysis. J Dermatolog Treat. 2016;27(4):316–21.

33. Wang J, Ke R, Shi W, Yan X, Wang Q, Zhang Q, et al. Association between psoriasis and asthma risk: A meta-analysis. Allergy Asthma Proc. 2018;39(2):103–9.

34. Hajiebrahimi M, Song C, Hägg D, Andersson TML, Villacorta R, Linder M. The Occurrence of Metabolic Risk Factors Stratified by Psoriasis Severity: A Swedish Population-Based Matched Cohort Study. Clin Epidemiol. 2020;12:737–44.

35. Blegvad C, Egeberg A, Tind Nielsen TE, Gislason GH, Zachariae C, Nybo Andersen AM, et al. Autoimmune Disease in Children and Adolescents with Psoriasis: A Cross-sectional Study in Denmark. Acta Derm Venereol. 2017;97(10):1225–9.

36. Ungprasert P, Raksasuk S. Psoriasis and risk of incident chronic kidney disease and end-stage renal disease: a systematic review and meta-analysis. Int Urol Nephrol. 2018;50(7):1277–83.

37. Egeberg A, Mallbris L, Hilmar Gislason G, Skov L, Riis Hansen P. Increased risk of migraine in patients with psoriasis: A Danish nationwide cohort study. J Am Acad Dermatol. 2015;73(5):829–35.

38. Hu SC, Lin CL, Tu HP. Association between psoriasis, psoriatic arthritis and gout: a nationwide population-based study. J Eur Acad Dermatol Venereol. 2019;33(3):560–7.

39. Patrick MT, Stuart PE, Raja K, Gudjonsson JE, Tejasvi T, Yang J, et al. Genetic signature to provide robust risk assessment of psoriatic arthritis development in psoriasis patients. Nature communications. 2018;9(1):4178.

40. Corbett M, Ramessur R, Marshall D, Acencio ML, Ostaszewski M, Barbosa IA, et al. Biomarkers of systemic treatment response in people with psoriasis: a scoping review. The British journal of dermatology. 2022;187(4):494–506.

