## Supplementary materials for "Body Composition, Cardiometabolic Risk Factors and Comorbidities in Psoriasis and the Effect of *HLA-C*06:02* Status: The HUNT Study, Norway"

**Supplementary material**

**Supplementary table 1. Demographic characteristics of the study population by sex.** All variables except sex, *HLA-C*06:02* frequency and disease duration are standardized for age.

|  | Psoriasis  (N= 3 535) | No psoriasis  (N= 49 838) |
| --- | --- | --- |
| Sex (N= 53 373) | | |
| Female, *n* (%) | 1 904 (53.9) | 27 125 (54.4) |
| Male, *n* (%) | 1 631 (46.1) | 22 713 (45.6) |
| Age, *years* (SD) (N = 53 373) | 57.3 (15.7) | 53.7 (17.6) |
| Female | 56.5 (16.2) | 53.2 (17.7) |
| Male | 58.3 (15.3) | 54.2 (17.3) |
| Smoking (N= 53 131) | | |
| Current daily, *n* (%) | 503.8 (13.5) | 4 091.4 (8.9) |
| Female | 338.9 (16.9) | 2543.1 (9.5) |
| Male | 166.2 (9.6) | 1554.9 (6.9) |
| No current daily, *n* (%) | 3 241.3 (86.6) | 45 300.5 (91.7) |
| Female | 1665 (83.1) | 24 338.4 (90.5) |
| Male | 1573.8 (90.5) | 20 956.1 (93.1) |
| Education (N = 53 024) | | |
| <10 years, *n* (%) | 521.4 (14.0) | 6 869.7 (13.9) |
| Female | 342.4 (16.2) | 4295.0 (16.0) |
| Male | 197.9 (11.4) | 2586.6 (11.5) |
| 10-12 years, *n* (%) | 1 905.0 (51.0) | 23 878.1 (48.5) |
| Female | 936.7 (46.9) | 11 824.4 (44.1) |
| Male | 966.2 (55.6) | 12 028.5 (53.5) |
| >12 years, *n* (%) | 1 310.2 (35.1) | 18 539.5 (37.6) |
| Female | 738.2 (36.9) | 10 688.4 (39.9) |
| Male | 572.7 (33.0) | 7864.1 (35.0) |
| *HLA-C*06:02* positive status, *n* (%) (N = 16 434) | 248 (26.6) | 1818 (11.7) |
| Female | 137 (27.7) | 974 (11.8) |
| Male | 111 (25.5) | 844 (11.6) |
| Psoriasis duration, *years* (SD) (N = 3 054) | 26.7 (17.6) | NA |
| Female | 26.3 (17.8) | NA |
| Male | 27.2 (17.3) | NA |

**Supplementary table II.**  **Body composition parameters in individuals with and without psoriasis by sex.** All variables are standardized for age.

|  |  | All participants  (N = 53 373) | | Psoriatic participants  (N= 901) | |
| --- | --- | --- | --- | --- | --- |
|  | Study group | Psoriasis  (N= 3 535) | No psoriasis  (N= 49 838) | *HLA-C*06:02-*positive  (N= 237) | *HLA-C*06:02-*negative  (N= 664) |
| Body composition parameters | | |  |  |  |
| Body mass index,  *kg/m^2^* (95% CI) | All participants | 28.3  (28.1, 28.4) | 27.3  (27.2, 27.3) | 27.9  (27.3, 28.5) | 28.5  (28.1, 28.9) |
|  | Females | 28.1  (27.9, 28.4) | 27.0  (26.9, 27.1) | 27.6  (26.8, 28.5) | 28.2  (27.5, 28.8) |
|  | Males | 28.4  (28.2, 28.6) | 27.6  (27.6, 27.7) | 28.1  (27.2, 29.0) | 28.9  (28.3, 29.4) |
| Waist circumference, *cm* (95% CI) | All participants | 100.6  (100.1, 101.1) | 97.4  (97.3, 97.5) | 100.1  (98.1, 102.1) | 102.0  (100.7, 103.2) |
|  | Females | 98.8  (97.4, 98.7) | 94.7  (94.5, 94.8) | 97.8  (95.1, 100.4) | 98.7  (97.0, 100.4) |
|  | Males | 103.5  (102.8, 104.2) | 100.6  (100.4, 100.8) | 102.7  (99.7, 105.8) | 105.1  (103.4, 106.8) |
| Percent body fat,  *%* (95% CI) | All participants | 33.3  (33.0, 33.6) | 31.5  (31.4, 31.6) | 31.9  (30.6, 33.1) | 32.5  (31.7, 33.2) |
|  | Females | 37.9  (37.5, 38.2) | 36.0  (35.9, 36.1) | 36.3  (34.9, 37.9) | 37.3  (36.4, 38.3) |
|  | Males | 28.1  (27.7, 28.5) | 26.3  (26.2, 26.4) | 26.9  (25.2, 28.5) | 27.8  (27.0, 28.7) |
| Visceral fat area,  *cm^2^* (95% CI) | All participants | 138.4 (136.4, 140.3) | 125.4  (125.0, 125.9) | 132.0  (124.0, 139.9) | 137.1  (132.4, 141.8) |
|  | Females | 150.6 (147.8, 153.3) | 136.0  (135.3, 136.7) | 143.7  (132.8, 154.7) | 148.8  (142.0, 155.7) |
|  | Males | 124.6 (122.0, 127.2) | 113.2  (112.6, 113.9) | 119.0  (107.8, 130.3) | 125.8  (119.7, 131.9) |
| Percent skeletal  muscle mass, *%* (95% CI) | All participants | 36.8  (36.6, 37.0) | 37.8  (37.8, 37.9) | 37.9  (37.1, 38.6) | 37.5  (37.1, 37.9) |
|  | Females | 33.8  (33.6, 34.0) | 34.8  (34.8, 34.9) | 34.9  (34.0, 35.7) | 34.3  (33.7, 34.8) |
|  | Males | 40.2  (40.0, 40.4) | 41.3  (41.2, 41.3) | 41.2  (40.2, 42.1) | 40.6  (40.1, 41.1) |
| Percent soft lean mass, *%* (95% CI) | All participants | 62.9  (62.6, 63.2) | 64.5  (64.4, 64.6) | 64.2  (62.9, 64.3) | 63.6  (62.9, 64.3) |
|  | Females | 58.5  (58.1, 58.8) | 60.2  (60.1, 60.3) | 59.8  (58.4, 61.3) | 59.0  (58.1, 59.8) |
|  | Males | 67.8  (67.4, 68.2) | 69.5  (69.4, 69.6) | 69.0  (67.4, 70.5) | 68.0  (67.2, 68.9) |
| Percent fat free mass,  % (95% CI) | All participants | 66.7  (66.4, 67.0) | 68.5  (68.4, 68.6) | 68.1  (66.9, 69.4) | 67.5  (66.8, 68.3) |
|  | Females | 62.1  (61.8, 62.5) | 64.0  (63.9, 64.1) | 63.6  (62.1, 65.1) | 62.7  (61.7, 63.6) |
|  | Males | 71.9  (71.5, 72.3) | 73.7 (73.6, 73.8) | 73.1  (71.5, 74.8) | 72.2  (71.3, 73.0) |
| Cardiovascular risk factors | | | | | |
| Systolic blood  pressure, *mmHg* (95% CI) | All participants | 132.0  (131.4, 132.6) | 131.1 (131.0, 131.3) | 127.3  (125.0, 129.7) | 127.1  (125.8, 128.4) |
|  | Females | 130.7  (129.8, 131.6) | 129.5 (129.3, 129.8) | 121.6  (118.4, 124.7) | 123.6  (121.8, 125.5) |
|  | Males | 133.5  (132.6, 134.3) | 133.0 (132.8, 133.3) | 133.5  (130.3, 136.7) | 130.5  (128.8, 132.3) |
| Diastolic blood pressure, *mmHg* (95% CI) | All participants | 74.1  (73.8, 74.4) | 74.0  (73.9, 74.1) | 74.6  (73.1, 76.0) | 73.5  (72.8, 74.3) |
|  | Females | 72.1  (71.6, 72.5) | 71.6  (71.5, 71.7) | 71.5  (69.7, 73.3) | 70.4  (69.5, 71.4) |
|  | Males | 76.4  (75.9, 76.9) | 76.8  (76.6, 75.9) | 77.9  (75.7, 80.0) | 76.5  (75.4, 77.7) |
| Total cholesterol, *mmol/L* (95% CI) | All participants | 5.4  (5.3, 5.4) | 5.3  (5.3, 5.3) | 5.2  (5.0, 5.4) | 5.3  (5.2, 5.4) |
|  | Females | 5.6  (5.5, 5.6) | 5.5  (5.5, 5.5) | 5.2  (5.1, 5.4) | 5.4  (5.3, 5.5) |
|  | Males | 5.1  (5.1, 5.2) | 5.2  (5.2, 5.2) | 5.2  (4.9, 5.4) | 5.2  (5.1, 5.3) |
| HDL-cholesterol, *mmol/L* (95% CI) | All participants | 1.4  (1.4, 1.4) | 1.4  (1.4, 1.4) | 1.3  (1.3, 1.4) | 1.3  (1.3, 1.4) |
|  | Females | 1.5  (1.5, 1.5) | 1.5  (1.5, 1.5) | 1.5  (1.4, 1.5) | 1.4  (1.4, 1.5) |
|  | Males | 1.2  (1.2, 1.2) | 1.3  (1.2, 1.3) | 1.2  (1.2, 1.3) | 1.2  (1.2, 1.2) |
| Triglycerides,  *mmol/L* (95% CI) | All participants | 1.8  (1.8, 1.8) | 1.7  (1.6, 1.7) | 1.8  (1.6, 2.0) | 1.9  (1.8, 2.0) |
|  | Females | 1.7  (1.6, 1.7) | 1.5  (1.5, 1.5) | 1.5  (1.4, 1.7) | 1.7  (1.5, 1.8) |
|  | Males | 1.9  (1.9, 2.0) | 1.8  (1.8, 1.8) | 2.1  (1.7, 2.4) | 2.1  (1.9, 2.2) |
| HbA1c, *mmol/L* (95% CI) | All participants | 36.4  (36.1, 36.7) | 35.3  (35.2, 35.3) | 34.1  (33.3, 34.8) | 34.6  (34.0, 35.1) |
|  | Females | 35.8  (35.4, 36.1) | 34.6  (34.5, 34.7) | 33.1  (32.4, 33.8) | 33.9  (33.1, 34.6) |
|  | Males | 37.1  (36.7, 37.5) | 36.1  (36.0, 36.2) | 35.1  (33.8, 36.4) | 35.3  (34.5, 36.0) |
| hsCRP, *mg/L* (95% CI) | All participants | 3.1  (2.9, 3.3) | 2.7  (2.7, 2.8) | 2.6  (2.2, 3.0) | 3.3  (2.9, 3.7) |
|  | Females | 3.3  (3.0, 3.6) | 2.9  (2.8, 3.0) | 2.6  (2.0, 3.2) | 3.7  (3.1, 4.3) |
|  | Males | 2.9  (2.6, 3.2) | 2.6  (2.5, 2.7) | 2.6  (1.9, 3.2) | 2.9  (2.3, 3.5) |

Abbreviations: kg, kilogram; cm, centimeter; SD, standard deviation; HDL-cholesterol, high-density lipoprotein cholesterol; HbA1c, glycated hemoglobin; hsCRP, high-sensitivity c-reactive protein

**Supplementary table III.** **Prevalence ratio (PR) for comorbid diseases in individuals with psoriasis compared to those without psoriasis, by sex.**

|  |  | **Psoriasis (N= 3535)** | **No psoriasis (N= 49 838)** | **Adjusted PR**  **(95% CI) ^a^** | **Fully adjusted PR (95% CI) ^b^** |
| --- | --- | --- | --- | --- | --- |
| **Cardiovascular disease** | | | | | |
|  | Myocardial infarction, *n* (%)  (N = 52 995) | 202 (6.0) | 1664 (3.3) | 1.54  (1.35, 1.76) | 1.44  (1.26, 1.66) |
|  | Female | 59 (3.2) | 427 (1.6) | 1.92 (1.48, 2.50) | 1.84 (1.41, 2.41) |
|  | Male | 143 (9.1) | 1217 (5.4) | 1.46 (1.24, 1.71) | 1.35 (1.15, 1.59) |
|  | Angina pectoris, *n* (%)  (N = 52 811) | 155 (4.6) | 1308 (2.7) | 1.55 (1.33, 1.82) | 1.53 (1.30, 1.80) |
|  | Female | 62 (3.4) | 480 (1.8) | 1.77 (1.37, 2.28) | 1.72 (1.31, 2.25) |
|  | Male | 93 (6.0) | 828 (3.68) | 1.43 (1.17, 1.75) | 1.42 (1.16, 1.74) |
|  | Heart failure, *n* (%)  (N = 52 779) | 90 (2.7) | 762 (1.5) | 1.56 (1.27, 1.93) | 1.54 (1.24, 1.92) |
|  | Female | 37 (2.0) | 298 (1.1) | 1.76 (1.26, 2.45) | 1.71 (1.20, 2.42) |
|  | Male | 53 (3.45) | 464 (2.1) | 1.47 (1.12, 1.93) | 1.46 (1.10, 1.93) |
|  | Atrial fibrillation, *n* (%)  (N = 52 486) | 259 (7.8) | 2430 (4.9) | 1.41 (1.25, 1.59) | 1.35 (1.19, 1.52) |
|  | Female | 108 (6.0) | 972 (3.6) | 1.52 (1.26, 1.88) | 1.48 (1.22, 1.80) |
|  | Male | 151 (9.8) | 1459 (6.5) | 1.34 (1.15, 1.56) | 1.27 (1.08, 1.49) |
|  | Apoplexia, *n* (%)  (N = 52 780) | 185 (5.5) | 1506 (3.1) | 1.63 (1.41, 1.88) | 1.56 (1.34, 1.81) |
|  | Female | 83 (4.6) | 692 (2.6) | 1.65 (1.33, 2.05) | 1.57 (1.25, 1.96) |
|  | Male | 102 (6.6) | 814 (3.6) | 1.61 (1.32, 1.95) | 1.55 (1.26, 1.90) |
| **Respiratory disease** | | | | | |
|  | Asthma, *n* (%)  (N = 52 915) | 552 (16.4) | 5612 (11.3) | 1.51 (1.40, 1.64) | 1.45 (1.34, 1.58) |
|  | Female | 313 (17.2) | 3150 (11.7) | 1.53 (1.38, 1.70) | 1.45 (1.30, 1.61) |
|  | Male | 239 (15.4) | 2462 (10.9) | 1.49 (1.32, 1.68) | 1.46 (1.29, 1.65) |
|  | Chronic obstructive pulmonary disease, *n* (%)  (N = 52 926) | 175 (5.2) | 1227 (2.5) | 1.89 (1.63, 2.20) | 1.72 (1.47, 2.01) |
|  | Female | 80 (4.4) | 591 (2.2) | 1.85 (1.48, 2.32) | 1.63 (1.29, 2.05) |
|  | Male | 95 (6.1) | 636 (2.8) | 1.92 (1.57, 2.36) | 1.79 (1.45, 2.21) |
| **Endocrine disease** | | | | | |
|  | Diabetes, *n* (%) (N = 53 243) | 336 (9.6) | 2699 (5.4) | 1.61 (1.45, 1.79) | 1.49 (1.31, 1.69) |
|  | Female | 154 (8.2) | 1251 (4.6) | 1.63 (1.39, 1.91) | 1.49 (1.23, 1.79) |
|  | Male | 182 (11.3) | 1448 (6.4) | 1.58 (1.37, 1.82) | 1.49 (1.25, 1.78) |
|  | Hypothyroidism, *n* (%)  (N = 52 499) | 293 (8.8) | 3072 (6.3) | 1.32 (1.18, 1.47) | 1.27 (1.14, 1.43) |
|  | Female | 230 (12.8) | 2496 (9.4) | 1.29 (1.14, 1.46) | 1.24 (1.10, 1.41) |
|  | Male | 63 (4.1) | 567 (2.6) | 1.44 (1.12, 1.86) | 1.39 (1.07, 1.80) |
|  | Hyperthyroidism, *n* (%)  (N = 52 275) | 98 (3.0) | 887 (1.8) | 1.58 (1.28, 1.93) | 1.54 (1.25, 1.89) |
|  | Female | 81 (4.6) | 728 (2.7) | 1.60 (1.28, 2.00) | 1.57 (1.25, 1.97) |
|  | Male | 17 (1.1) | 159 (0.7) | 1.45 (0.88, 2.39) | 1.37 (0.82, 2.29) |
| **Others** | | | | | |
|  | Migraine, *n* (%) (N = 52 930) | 707 (21.0) | 7622 (15.4) | 1.41 (1.31, 1.50) | 1.39 (1.30, 1.49) |
|  | Female | 491 (27.0) | 5641 (20.9) | 1.33 (1.23, 1.44) | 1.32 (1.22, 1.43) |
|  | Male | 216 (14.0) | 1980 (8.8) | 1.67 (1.46, 1.90) | 1.63 (1.42, 1.87) |
|  | Renal disease (Non-UTI), *n* (%) (N = 52 909) | 188 (5.6) | 1584 (3.2) | 1.63 (1.41, 1.89) | 1.53 (1.31, 1.78) |
|  | Female | 101 (5.6) | 928 (3.4) | 1.54 (1.26, 1.88) | 1.44 (1.16, 1.77) |
|  | Male | 87 (5.6) | 656 (2.9) | 1.67 (1.46, 1.90) | 1.63 (1.30, 2.05) |
|  | Gout, *n* (%) (N = 52 791) | 246 (7.3) | 1778 (3.6) | 1.83  (1.61, 2.07) | 1.63 (1.44, 1.85) |
|  | Female | 91 (5.1) | 581 (2.2) | 2.16 (1.74, 2.67) | 1.82 (1.46, 2.27) |
|  | Male | 155 (10.0) | 1197 (5.3) | 1.69 (1.45, 1.98) | 1.56 (1.33, 1.83) |

^a^ Adjusted for age and sex, except in sex-specific analyses in which sex is excluded as a covariate

^b^ Adjusted for age, sex, BMI, education and smoking, except in sex-specific analyses in which sex is excluded as a covariate

Abbreviations: CI, confidence interval; UTI, urinary tract infection; BMI, body mass index

**Supplementary table IV.** **Prevalence ratio (PR) for comorbid diseases in *HLA-C*06:02*-positive compared with *HLA-C*06:02*-negative individuals with psoriasis, by sex.**

|  | Non-related psoriatic participants (N= 901) | | |
| --- | --- | --- | --- |
|  | *HLA-C*06:02*  -positive  (N = 248) | *HLA-C*06:02*  -negative  (N = 683) | Adjusted PR (95% CI) ^a^ |
| Cardiovascular disease |  |  |  |
| Myocardial infarction, *n* (%) (N = 898) | 6 (2.7) | 18 (2.8) | 1.09 (0.37, 3.19) |
| Female | 3 (2.4) | 2 (0.6) | 17.04 (1.33, 219.05) |
| Male | 3 (3.0) | 16 (5.2) | 0.75 (0.18, 3.19) |
| Angina pectoris, *n* (%) (N = 891) | 11 (4.9) | 19 (3.0) | 1.39 (0.59, 3.27) |
| Female | 7 (5.6) | 6 (1.8) | 2.83 (0.74, 10.78) |
| Male | 4 (4.0) | 13 (4.3) | 1.05 (0.29, 3.81) |
| Heart failure, *n* (%) (N = 891) | 3 (1.3) | 6 (0.9) | 2.06 (0.15, 28.55) |
| Female | 2 (2.4) | 1 (0.30) | 22.27 (3.26, 152.39) |
| Male | 0 (0.0) | 5 (1.7) | NA |
| Atrial fibrillation, *n* (%) (N = 884) | 18 (8.0) | 26 (4.1) | 3.69 (1.85, 7.35) |
| Female | 11 (8.7) | 6 (1.8) | 4.82 (1.35, 17.18) |
| Male | 7 (7.0) | 20 (6.6) | 2.50 (1.13, 5.51) |
| Apoplexia, *n* (%) (N = 890) | 7 (3.1) | 21 (3.3) | 0.84 (0.28, 2.54) |
| Female | 5 (4.0) | 10 (3.0) | 1.47 (0.37, 5.89) |
| Male | 2 (2.0) | 11 (3.6) | 0.35 (0.05, 2.61) |
| Respiratory disease |  |  |  |
| Asthma, *n* (%) (N = 890) | 31 (13.8) | 116 (18.2) | 0.80 (0.53, 1.21) |
| Female | 20 (16.1) | 63 (19.0) | 0.80 (0.46, 1.39) |
| Male | 11 (10.9) | 53 (17.4) | 0.73 (0.39, 1.36) |
| COPD, *n* (%) (N = 890) | 7 (3.1) | 18 (2.8) | 1.23 (0.33, 4.51) |
| Female | 3 (2.4) | 8 (2.4) | 1.38 (0.20, 9.70) |
| Male | 4 (3.9) | 10 (3.3) | 1.58 (0.24, 10.57) |
| Endocrine disease |  |  |  |
| Diabetes, *n* (%) (N = 918) | 11 (4.8) | 41 (6.2) | 0.75 (0.34, 1.67) |
| Female | 5 (4.0) | 19 (5.5) | 0.62 (0.18, 2.15) |
| Male | 6 (5.7) | 22 (7.0) | 1.02 (0.40, 2.62) |
| Hypothyroidism, *n* (%) (N = 884) | 12 (5.4) | 41 (6.5) | 0.94 (0.48, 1.87) |
| Female | 10 (8.0) | 35 (10.5) | 1.00 (0.51, 1.97) |
| Male | 2 (2.0) | 6 (2.0) | 0.70 (0.08, 6.10) |
| Hyperthyroidism, *n* (%) (N = 878) | 5 (2.3) | 12 (1.9) | 1.24 (0.39, 3.99) |
| Female | 4 (3.3) | 11 (3.3) | 1.00 (0.27, 3.65) |
| Male | 1 (1.0) | 1 (0.3) | NA |
| Others |  |  |  |
| Migraine, *n* (%) (N = 888) | 50 (22.3) | 179 (28.1) | 0.73 (0.54, 0.98) |
| Female | 38 (30.4) | 120 (35.8) | 0.83 (0.60, 1.16) |
| Male | 12 (12.1) | 59 (19.6) | 0.52 (0.28, 0.99) |
| Renal disease (Non-UTI), *n* (%) (N = 891) | 6 (2.7) | 27 (4.2) | 0.42 (0.13, 1.42) |
| Female | 3 (2.4) | 14 (4.2) | 0.37 (0.08, 1.62) |
| Male | 3 (3.0) | 13 (4.7) | 0.47 (0.06, 3.88) |
| Gout, *n* (%) (N = 888) | 10 (4.5) | 40 (6.3) | 1.17 (0.57, 2.40) |
| Female | 6 (4.92) | 17 (5.1) | 1.62 (0.59, 4.46) |
| Male | 4 (4.0) | 23 (7.6) | 0.78 (0.27, 2.24) |

^a^ Adjusted for age, sex, BMI, smoking status, disease duration and cryptic population structure by four principal components (PCs), except sex-specific analyses for which sex was excluded as a covariate.

Abbreviations: CI, confidence interval; UTI, urinary tract infection; BMI, body mass index

**Supplementary figures**

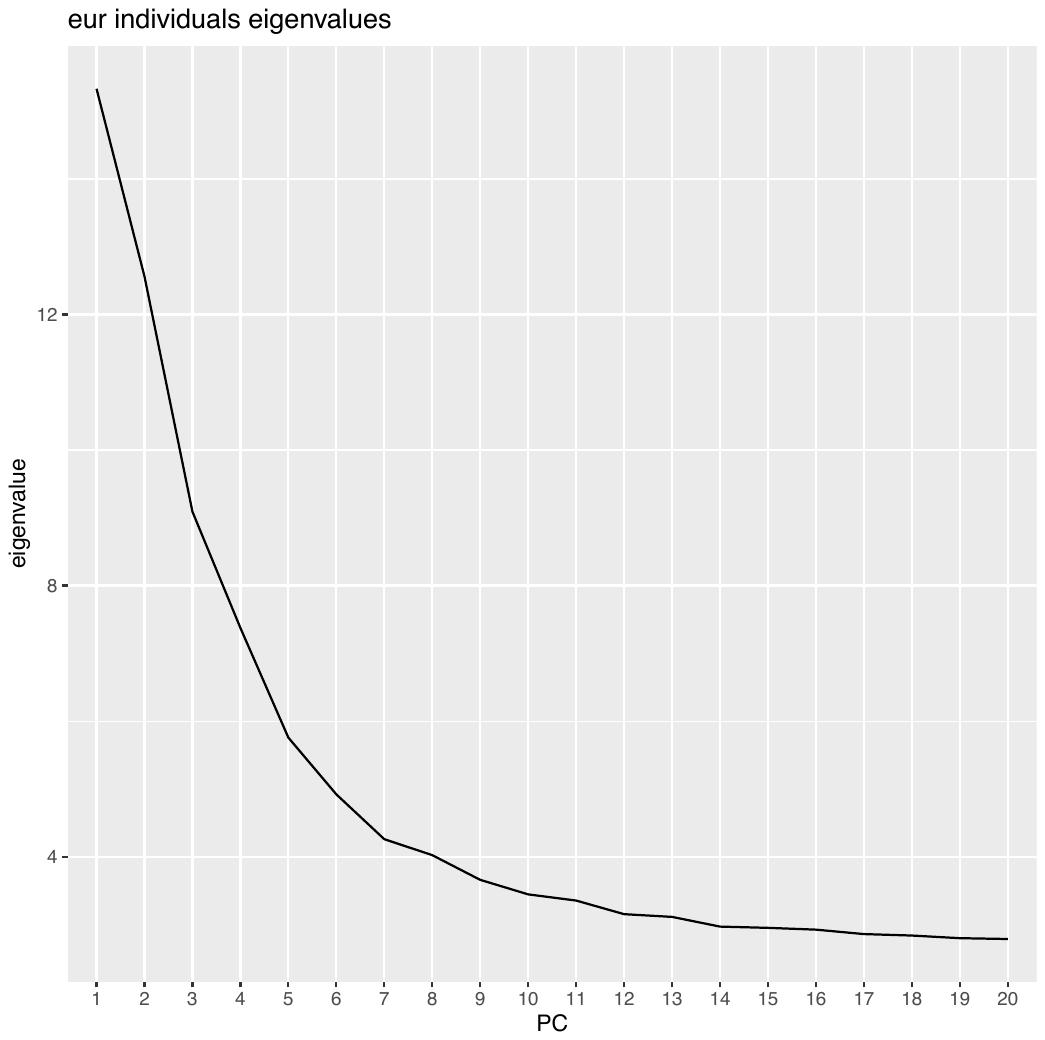

**Supplementary figure 1:** Eigen values for each principal component in individuals of European descent

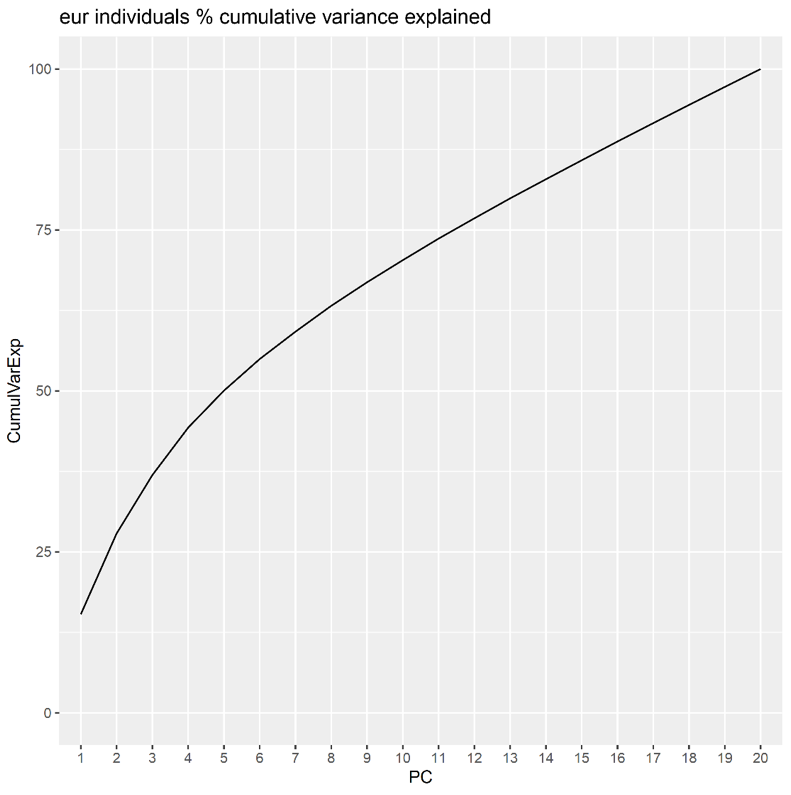

**Supplementary figure 2:** Cumulative variance explained by the first N components in individuals of European decent
